# Nowcasting Influenza-like Illness Trends in Cameroon

**DOI:** 10.1101/2020.07.02.20145250

**Authors:** Elaine O. Nsoesie, Olubusola Oladeji, Aristide S. Abah Abah, Martial L. Ndeffo-Mbah

## Abstract

Although acute respiratory infections are a leading cause of mortality in sub-Saharan Africa, surveillance of diseases such as influenza is mostly neglected. Evaluating the usefulness of influenza-like illness (ILI) surveillance systems and developing approaches for forecasting future trends is important for pandemic preparedness. We applied statistical and machine learning models to forecast 2012 to 2018 trends in ILI cases reported by the Cameroon Ministry of Health (MOH), using Google searches for influenza symptoms, treatments, natural or traditional remedies as well as, infectious diseases with a high burden (i.e., AIDS, malaria, tuberculosis). The variance explained by the models based on Google search data were 87.7%, 79.1% and 52.0% for the whole country, the Littoral and Centre regions respectively. Our study demonstrates the need for developing contextualized approaches when using digital data for disease surveillance and demonstrates the potential usefulness of search data for monitoring ILI in sub-Saharan African countries.

## INTRODUCTION

Influenza and other respiratory tract infections remain a global public health issue in spite of vaccine availability^1^. Every year, there are about 650,000 deaths globally and three to five million respiratory illnesses due to the influenza virus^2^. In Africa, acute respiratory infections are thought to be a major cause of morbidity and mortality^3,4^, despite considerable under-reporting and sparsity of precise assessments of the total health and economic burden across the continent ^3,5^.

The 2009 influenza pandemic reinforced the necessity for developing robust influenza surveillance systems globally ^6^. Studies published right after the pandemic noted that many countries in Africa were not equipped with sufficient data on the epidemiology and risk factors of influenza to put in place the necessary approaches for influenza prevention and control^7,8^. This gap in global early-detection and response measures has motivated extensive research towards improving influenza surveillance using both traditional and non-traditional data streams ^9–19^, as well as the development of a global standard for influenza surveillance by the World Health Organization (WHO)^20^. Though influenza surveillance data from Africa has improved in recent years, data quality is still lagging behind those implemented in other regions of the world. This lagging is mainly due to limited capacity for laboratory confirmation of influenza diagnoses and timely reporting of virological and epidemiological data. From a global perspective, understanding influenza dynamics in the tropics is important for pandemic planning and response since poor surveillance capabilities can delay the detection of novel viruses^7,21^. Although more than a billion people are living in tropical regions, there are no sufficient data on influenza-related mortality and morbidity ^21^.

Digital epidemiology has enabled the use of non-traditional data sources (e.g., social media, Internet search queries) to monitor the spread of influenza-like illness (ILI) ^9,10,16,22–25^. However, to our knowledge, no studies have focused on applying digital epidemiology to ILI surveillance in a sub-Saharan African country using data collected by a national surveillance system. Here, we seek to demonstrate that the adoption of digital data for public health surveillance in African countries requires a contextualized approach^26,27^. For example, cultural and societal practices unique to the sub-Saharan African context demands a rethinking of what search terms should be included in models used for surveillance. An individual with influenza-like symptoms might search for terms such as, “cold” or “catarrh”, but might also search for traditional or natural remedies and diseases such as HIV, TB and malaria, which have a high burden across sub-Saharan Africa and are well-known.

In this paper we aim to model and nowcast (hereafter referred to as, forecast) ILI in Cameroon using data from a surveillance system for influenza and ILI implemented by the Ministry of Health (MOH) in Cameroon^28^. Recent studies on influenza and ILI surveillance in Cameroon have focused on viral identification ^29^, understanding genetic diversity^30,31^, and seasonality of circulating viruses ^32^. Here, we focus on providing a framework for forecasting future trends, which could be invaluable for public health planning and response.

## RESULTS

The number of reported ILI cases varied across the ten regions in Cameroon. The neighboring East and Adamaoua regions (SI Figure 1) had the highest number of reported cases; 17.1 (95% CI, 15.9 - 18.3) and 16.4 (95% CI, 15.7 - 17.1) reports per 100,000 people, respectively (Figure 1). The major economic regions, Littoral and Centre regions which had the most consistent reporting, had different attack rates: 8.3 (95% CI, 7.8 - 8.9) and 12.6 (95% CI, 12.1 - 13.1), respectively. There were no distinct regional groupings in reporting patterns.

**Figure 1:**
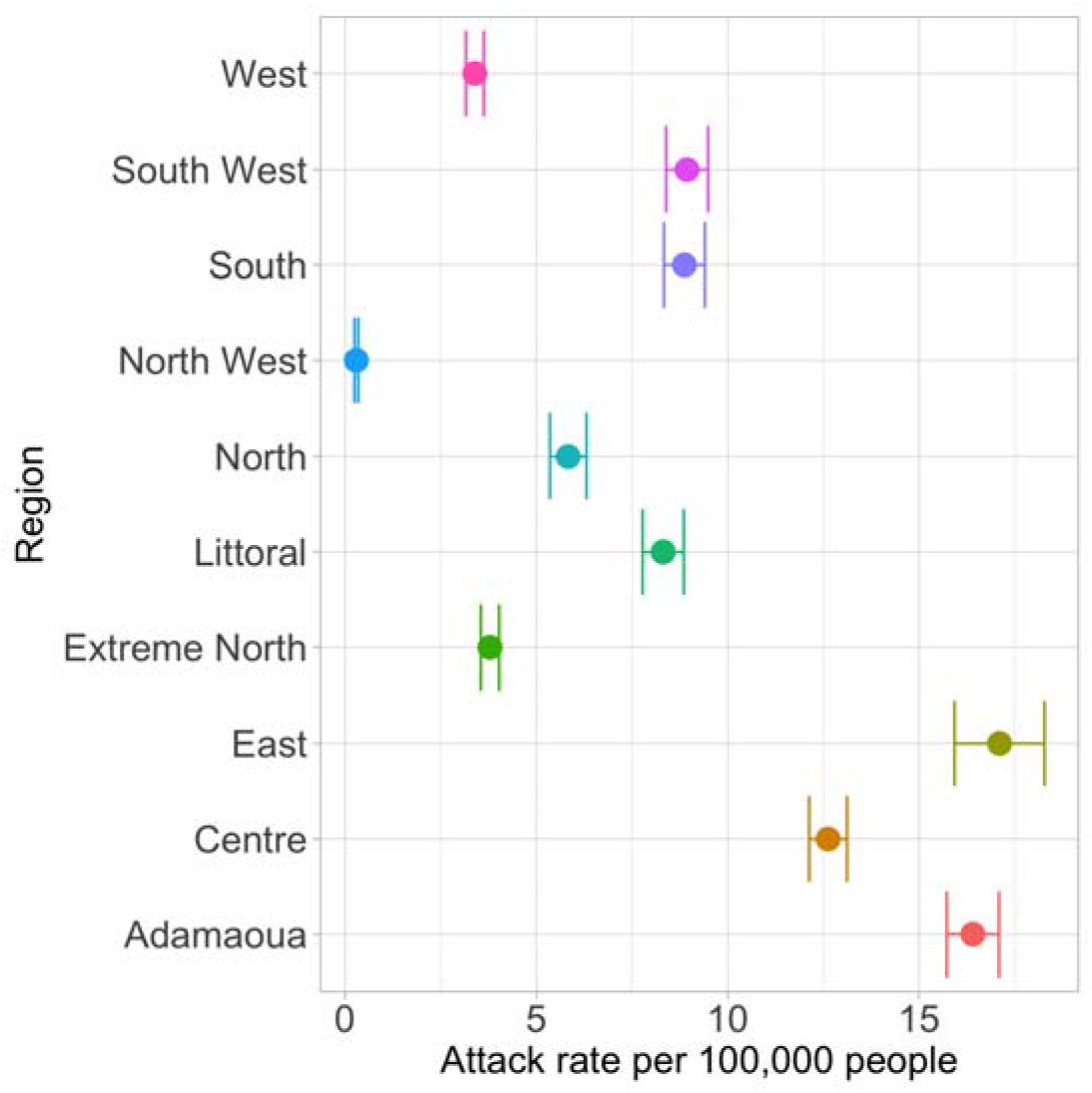
Distribution of ILI reports across the timespan of the study (mean and 95% confidence interval).

### Out-of-sample Predictions

Out-of-sample predictions based on the leave one year out approach described in the methods produced varying results across the regions. At the national level, search terms (SI Table 1) that had correlations of 0.3 or greater with ILI were used as explanatory variables in the models. These included, catarrh, pain, douleur (pain), transpire (sweat), gingembre (ginger), miel (honey), tea, malaria, paludisme (malaria), TB, AIDS, and HIV. SVM had the lowest mean RMSE and the highest mean *R*^*2*^ — 1.078 and 0.877, respectively (Figure 2a & b). Random Forest had a mean RMSE of 1.44 and *R*^*2*^ of 0.771. The models that did not include search terms had comparable RMSE and *R*^*2*^ to Random Forest. However, the corresponding RMSE range for out-of-sample predictions using SVM was wider when compared to the other models (Figure 2c); suggesting that SVM might not perform well in long range forecasts.

**Figure 2:**
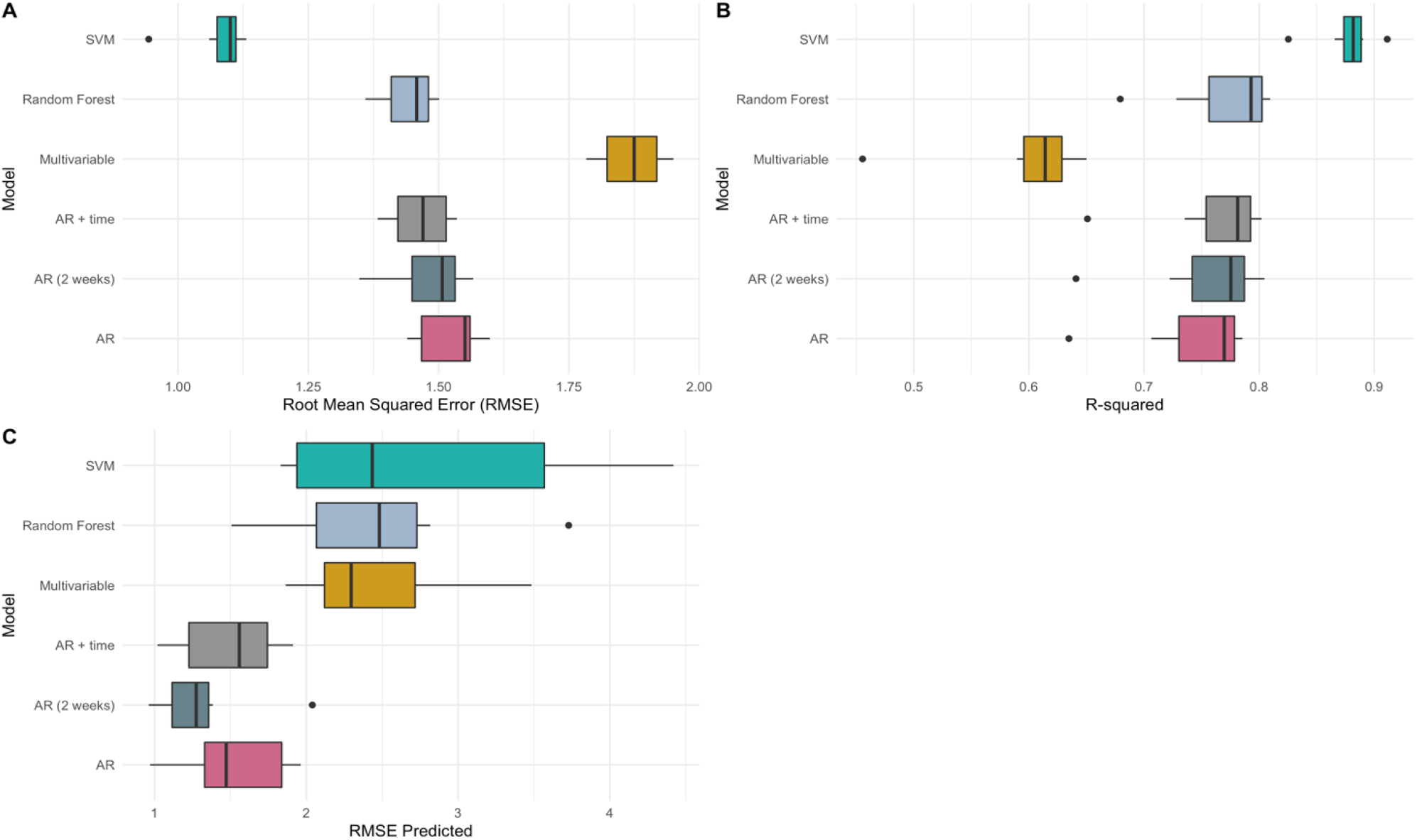
Comparison of models at the national level. (a) Root Mean Squared Error (RMSE) of fitted values, (b) model R-squared and (c) RMSE of out-of-sample predictions. The models are Support Vector Machines Regression (SVM), Random Forest Regression, Multivariable linear regression, Autoregressive model with time and ILI values at time t-1 included as explanatory variables (AR + time), Autoregressive model (AR) based on ILI at time t-1, and Autoregressive model (AR (2 weeks)) that includes ILI data at time t-1 and t-2 as explanatory variables.

The regional models for Littoral performed similarly to the national models. The mean RMSE for SVM and Random Forest were 2.412 and 2.942, respectively (Figure 3a). While the corresponding average *R*^*2*^ were 0.791 and 0.677 (Figure 3b). The mean *R*^*2*^ for the autoregressive linear regression models were 0.675, 0.690, and 0.707 for the AR, AR (2 weeks) and AR + time models, respectively. The respective mean RMSE were also in a similar range as the models that included search terms — 2.931, 2.855, and 2.784. The mean out-of-sample RMSE (Figure 3c) values were also similar, however, the ranges for models with search terms as explanatory variables were broader. The search terms included in all the models were rhume (cold), cough, douleur (pain), aloe vera, citron (lemon), gingembre (ginger), miel (honey), malaria, TB, AIDS and HIV.

**Figure 3:**
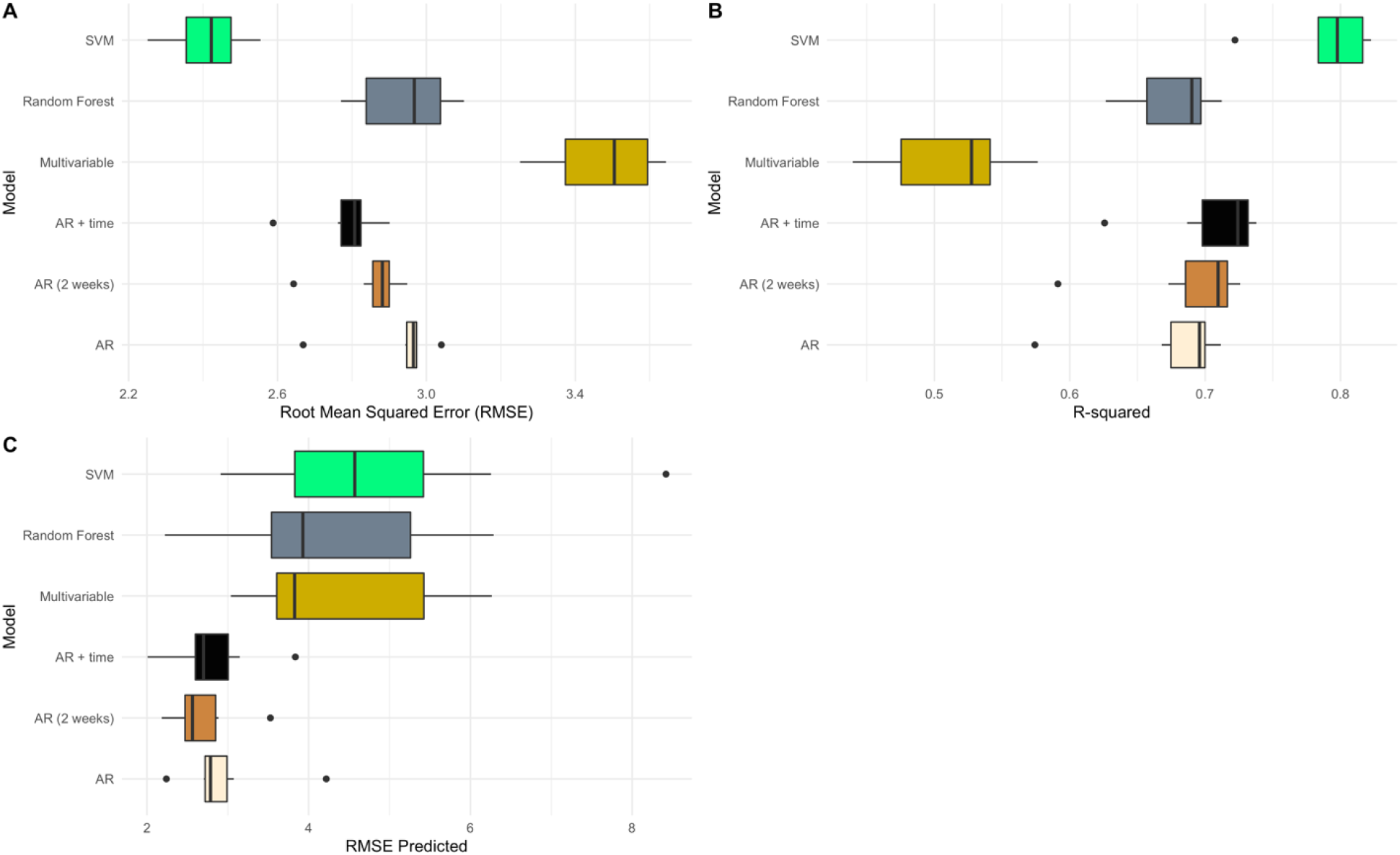
Comparison of models fitted to data for the Littoral region. (a) Root Mean Squared Error (RMSE) of fitted values, (b) model R-squared and (c) RMSE of out-of-sample predictions. The models are Support Vector Machines Regression (SVM), Random Forest Regression, Multivariable linear regression, Autoregressive model with time and ILI values at time t-1 included as explanatory variables (AR + time), Autoregressive model (AR) based on ILI at time t-1, and Autoregressive model (AR (2 weeks)) that includes ILI data at time t-1 and t-2 as explanatory variables.

In contrast to the Littoral and national models, only three search terms were included in models for the Centre region; gingembre (ginger), lemon and AIDS. The data was noisier, the RMSE was higher for all models (Figure 4a) and the highest variance explained was 52.0%; much lower compared to the national and Littoral models (Figure 4b). The autoregressive models performed similarly, while the multivariable linear regression models had the poorest goodness of fit (mean *R*^*2*^ = 0.190) (Figure 4c). These differences across the three regions show variability in data quality and highlight the need for regional models that capture underlying trends in reporting and data quality.

**Figure 4:**
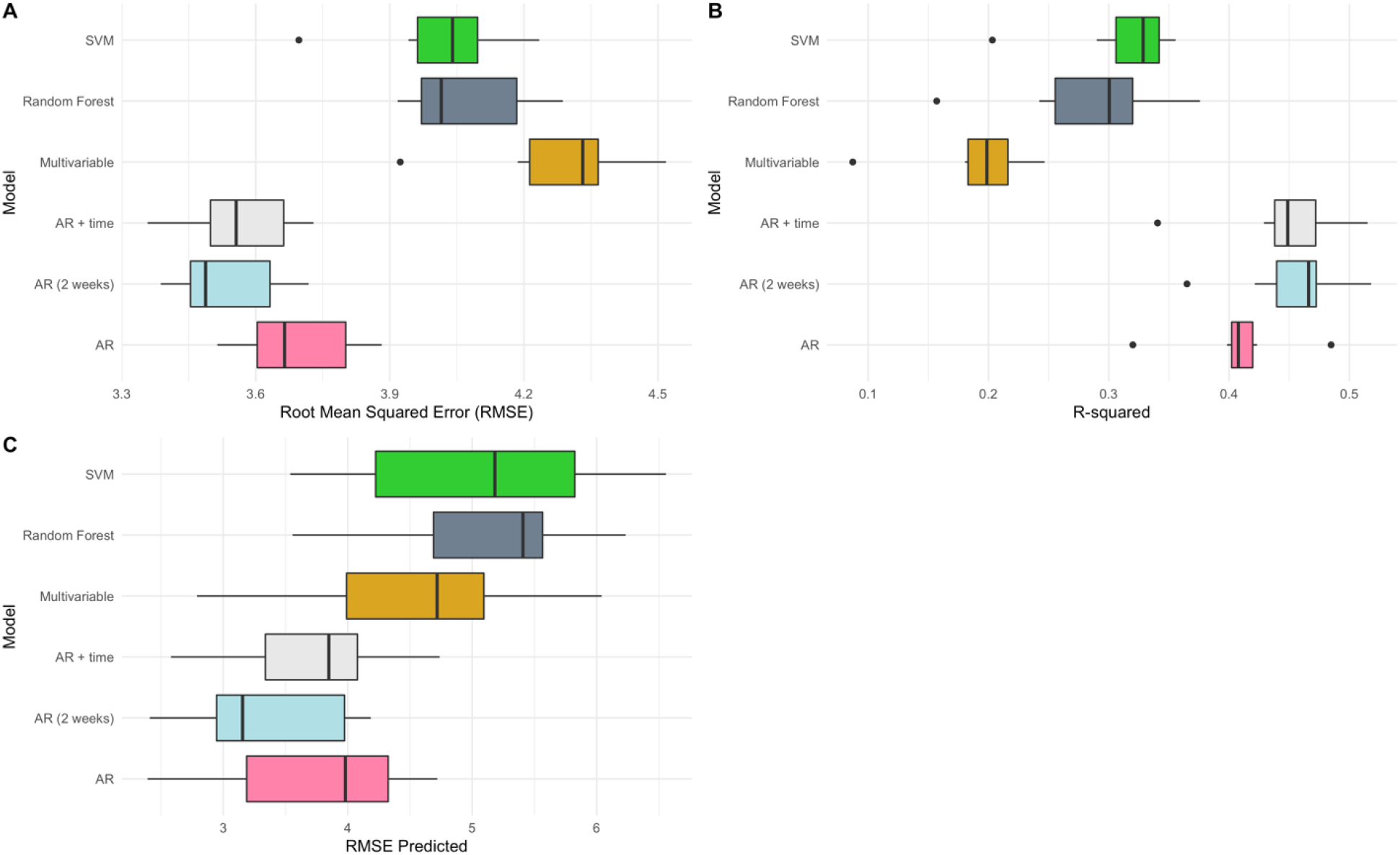
Comparison of models fitted to data for the Centre region. (a) Root Mean Squared Error (RMSE) of fitted values, (b) model R-squared and (c) RMSE of out-of-sample predictions. The models are Support Vector Machines Regression (SVM), Random Forest Regression, Multivariable linear regression, Autoregressive model with time and ILI values at time t-1 included as explanatory variables (AR + time), Autoregressive model (AR) based on ILI at time t-1, and Autoregressive model (AR (2 weeks)) that includes ILI data at time t-1 and t-2 as explanatory variables.

### One-week-ahead Forecasts

The correlation between the national percent ILI and forecasted values were 72.9% and 66.8% using the SVM and Random Forest models, respectively (Figure 5). The search terms by order of importance in the Random Forest model were as follows: AIDS, catarrh, HIV, paludisme, gingembre, tea, miel (honey), malaria, transpire (sweat), douleur (pain), TB, and pain. Similar results were obtained for the Littoral region, where the correlation between the predicted ILI values and the actual percent ILI was 80.9% and 60.29% for SVM and Random Forest, respectively (Figure 6). While the point estimates using the SVM model were more aligned with the actual ILI values, the confidence intervals for Random Forest were more robust. In contrast, the best forecast for the Centre region was based on the autoregressive model using solely ILI data from the last two time points. The correlation between the forecasted values and the ILI data was 44.6%.

**Figure 5:**
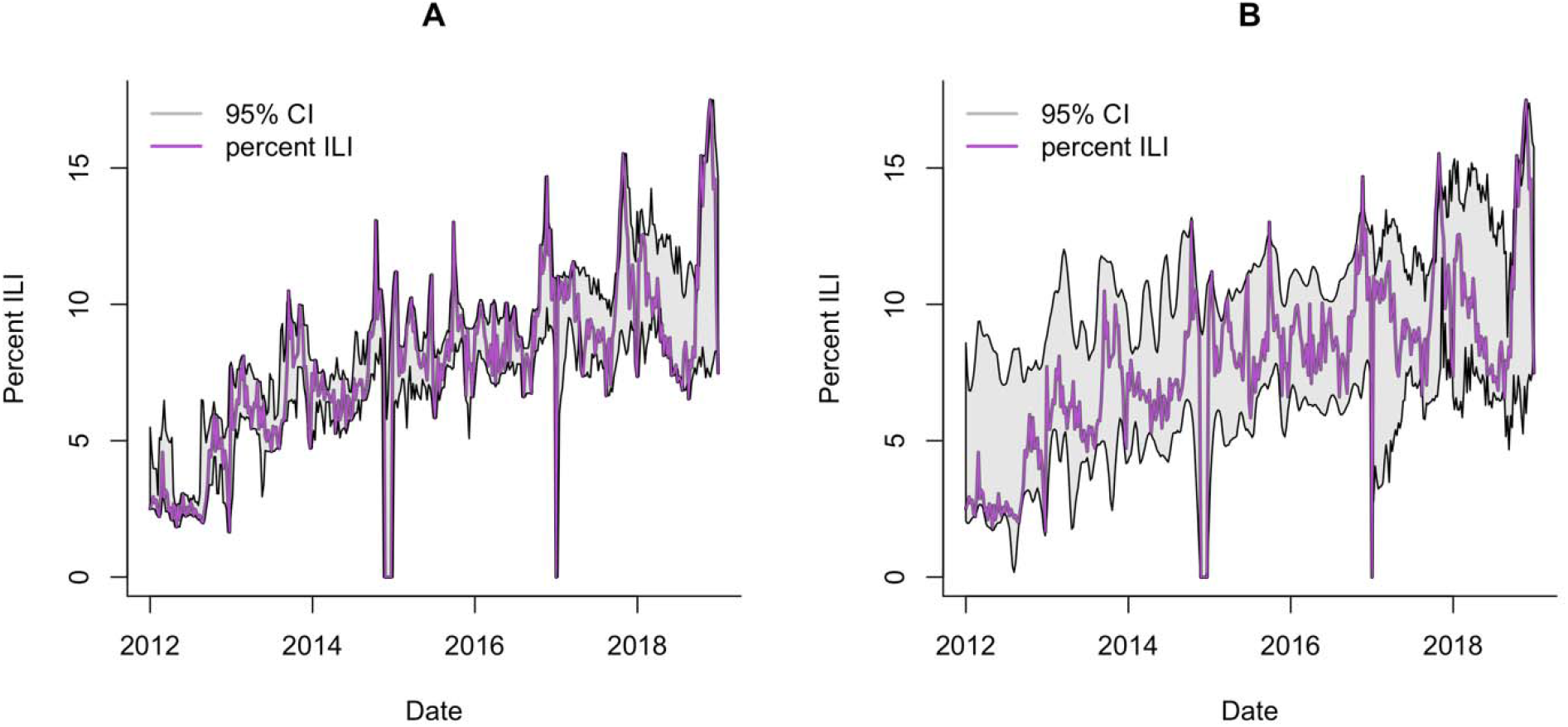
95% Confidence Intervals (CI) for one-week-ahead predictions of percent ILI at the national level. Models were fit using five years of data; week 1 to 261. Forecasts for the next week, t+1, were made at week, t. Estimates using (a) Random Forest Regression and (b) Support Vector Machine Regression (SVM) are presented.

**Figure 6:**
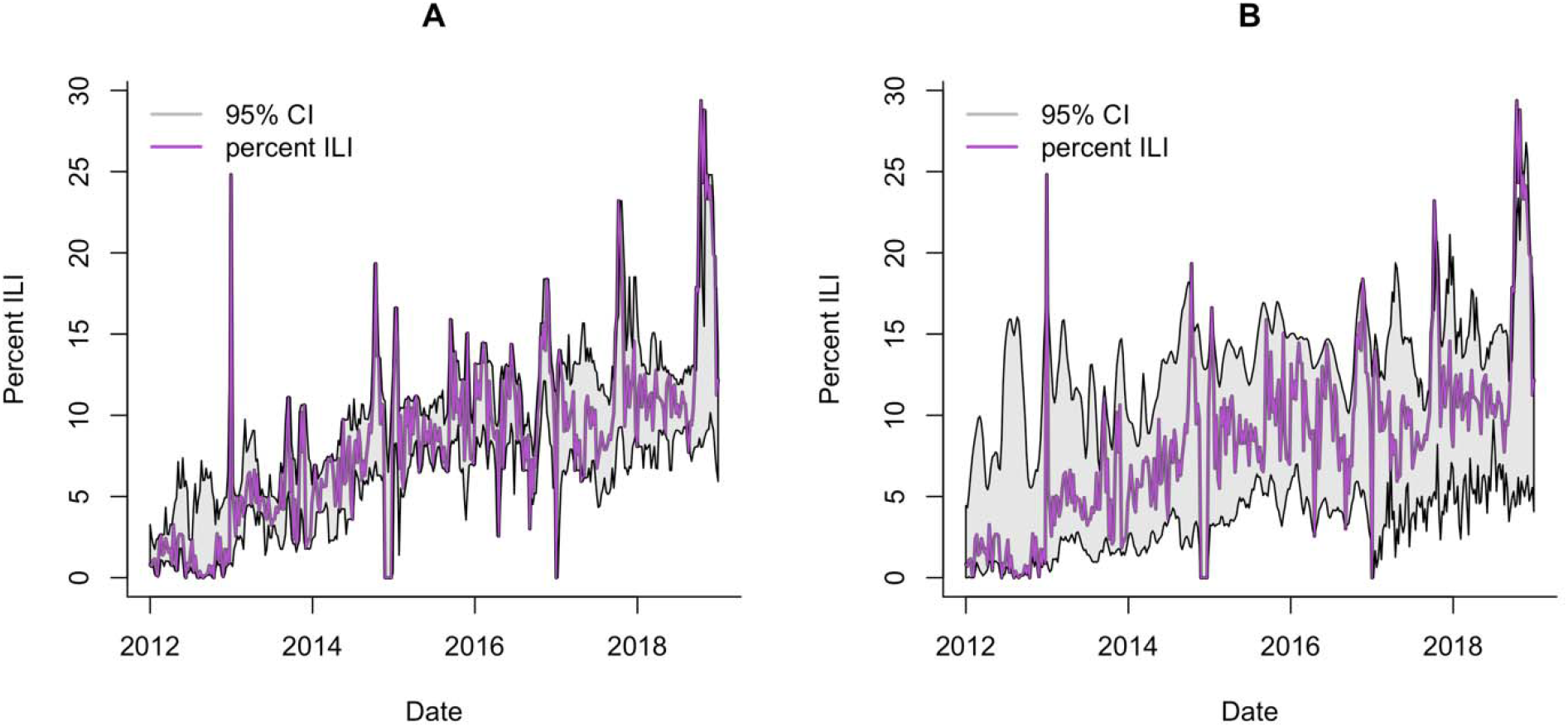
95% Confidence Intervals (CI) for one-week-ahead predictions of percent ILI for the Littoral region. Models were fit using five years of data; week 1 to 261. Forecasts for the next week, t+1, were made at week, t. Estimates using (a) Random Forest Regression and (b) Support Vector Machine Regression (SVM) are presented.

## DISCUSSION

Influenza causes considerable disease burden in tropical and subtropical regions. However, despite its global significance, influenza surveillance systems in sub-Saharan Africa are lagging behind those in other regions of the world. National surveillance systems can fill in this gap in monitoring influenza-like illnesses, including emerging viruses, if the systems are well-designed, maintained and regularly evaluated. However, as noted across several countries including the United States, ILI reports from official sources such as, the Centers for Disease Control and Prevention (CDC) can be delayed ^9,33^. Digital data sources can be used to supplement data from official sources.

In this study, we highlight three important findings. First, health information seeking trends on the Internet are useful for monitoring health information needs in sub-Saharan African countries^27^. Second, as mobile phone and Internet usage increases across the continent, data produced using these technologies can provide a supplementary approach for monitoring disease trends. However, the local context is important in both research and interpretation of findings. Search terms commonly associated with ILI in western countries might not be associated with ILI in sub-Saharan African countries. Third, our study also highlights a need for public health education about influenza and influenza-like illness in Cameroon. Africans, like people in other regions of the world, use the Internet for information seeking. Understanding how different Internet platforms are used can be useful for designing appropriate public health interventions, including disease prevention measures and education on the causes and treatment of diseases^27^.

Our findings suggest that combining search data related to influenza symptoms, treatments, natural or traditional remedies as well as infectious diseases with a high burden (i.e., AIDS, malaria, tuberculosis) in sub-Saharan countries could be useful for tracking ILI trends. Specifically, terms such as, cold, catarrh, and pain, and home remedies such as, tea, ginger and honey, were important predictors of ILI trends. Searches for AIDS and HIV might be due to HIV confirmed individuals searching if ILI symptoms are considered HIV symptoms. Studies have shown that individuals prefer using search engines to seek information on stigmatized topics ^34^.

Also, our methods perform best in short-term forecasting, but also suggest some usability for long-term forecasting as demonstrated in the leave one year out model evaluation. Furthermore, autoregressive models using previous values of ILI to predict current values achieved reasonable accuracy suggesting that previously observed ILI trends can be used for predicting current and future trends. The machine learning regression methods, Random Forest and Support Vector Machines, performed similarly with some minor differences. The confidence intervals in the SVM model were more heavily influenced by variability in the search data and the ILI reports, when compared to the Random Forest estimates. Future studies will focus on developing ensemble approaches involving the consensus of multiple algorithms for real-time forecasting.

Syndromic and pre-diagnostic healthcare data (e.g., school absenteeism, pharmacy sales) in conjunction with statistical methods to monitor and characterize epidemics earlier than traditional public health systems and to provide situational awareness for ongoing outbreaks has been shown to be useful ^35,36^. However, clear definitions of disease symptoms are important, especially for regions such as sub-Saharan Africa where many diseases can cause influenza-like symptoms. To accomplish this, interventions including education on the causes and treatments of influenza-like disease are needed. These non-traditional methods can supplement traditional surveillance, however, the need for developing reliable surveillance systems remains. Additionally, consistent data collection procedures and case definition are necessary for evaluation of ILI surveillance systems. ILI cases are most likely severely underreported in Cameroon, and there is not sufficient information to adjust the data accordingly.

Another challenge with forecasting ILI trends in subtropical and tropical regions is that seasonality is less defined; some regions experience semi-annual epidemics or annual influenza activity without a well-defined season while others have yearly epidemics that correspond with the rainy season ^17,37^. Indoor crowding during the rainy season has been suggested as a possible driver of influenza epidemics in the tropics ^21,38^. In contrast, in the temperate regions of the world, influenza has marked seasonal patterns with increased rates during the winter months, followed by low activity during the warmer months ^39,40^.

Despite these limitations, these data have the potential to improve the timely detection of outbreaks and surges in unusual disease activity requiring further public health investigation. A lack of comprehensive health data in most sub-Saharan African countries is a major roadblock to disease control. The most effective approach to mitigating a potential pandemic is to stem disease emergence at its location of origin, the success of which depends on efficient disease surveillance systems. Addressing gaps in surveillance in sub-Saharan Africa should be a priority in our global preparedness and response strategies for influenza and other acute respiratory infections. Data from digital sources could support the development of up-to-date disease surveillance systems.

## METHODS

### ILI Data

The Cameroon sentinel surveillance system aims to detect the current type of influenza viruses and involves twenty-two health facilities across the country. The system also involves influenza-like illness surveillance based on symptom definition and is designed for early warning detections of surges. ILI was defined as sudden onset of fever (temperature >38°C) and cough or sore throat, with the onset of symptoms within the five days before presentation at a health facility based on the WHO’s Integrated Disease Surveillance and Response (IDSR) ^41^. The MOH provided a weekly number of cases reported each week to the Cameroon Ministry of Health (MOH) in Yaoundé from January 2012 to December 2018.

### Google Search Data

We obtained the weekly volume of Google searches for 65 terms (see SI Table1) in both French and English from the Google API at the national level, and for the Centre and Littoral regions from January 2012 to December 2018. Data was sparser for the other regions. The Google search terms consisted of ILI symptoms, home remedies typically used in Cameroon for colds, and inquiries for malaria, HIV/AIDs, and TB. We were particularly interested in home remedies because it captures a context-specific approach to managing ILI symptoms. Next, we excluded all search terms that had zero variance or had a Pearson correlation of less than 0.3, with weekly ILI reports from our data so we could focus our analysis on the most relevant terms. This resulted in three, eleven and twelve terms separately for the Centre region, Littoral region and the whole country.

### Statistical Analysis

To assess the potential usefulness of our approach for public health decision making in Cameroon, we performed two sets of analysis. First, we used a cross-validation approach whereby at each modeling step, one year of data is excluded from the fitting and used in the model validation. Since there are seven years of data, this implies seven different models were fitted. Next, the best modeling approach for the Centre and Littoral regions, and the whole country based on the model R-squared and the root mean squared error (RMSE) were used in one-week ahead forecasting. This allows for forecasting the current ILI percentage.

We evaluated several multivariable regression models with the search data as explanatory variables and the ILI as the dependent variable. Regression models are traditional statistical models that are widely used to analyze quantitative data and make predictions about future values of the data. Here, we considered a range of robust regression modeling approaches that have been previously used for forecasting influenza trends ^11,12,42^: random forest regression, support vector regression, multivariable linear regression and autoregressive time series regression models.

Random Forest is an ensemble of regression trees created by using bootstrap samples of the data and random feature selection in tree induction. To construct regression trees each node is split using the best split among a subset of predictors randomly chosen at that node. It is an extension of bagging – bootstrap aggregating – a method for combining several predictors to decrease the variance of the prediction function ^43–45^. There are several advantages to random forests including, high accuracy, robustness to overfitting, and estimation of important variables ^45^. These are useful for identifying which covariates are most significant in the model ^45^. Regression with Support Vector Machines (SVM) involves mapping the independent variables into a high-dimensional feature space using a linear or non-linear kernel function^46^. SVM regression with the linear kernel is akin to multivariate linear regression models. We used a radial kernel in SVM, since it fitted best to the data trend, and was selected using the tuning function in R ^47,48^. The random forest, SVM, and multivariable linear regression models involved fitting ILI to the Google search terms as explanatory variables. To capture the potential delay between the time when an individual with symptoms searches for information online, and when they visit a healthcare facility, we lagged some search terms for up to four weeks in the regression models based on prior examination of the cross correlations between the ILI data and search terms. We also applied LOESS (Local Polynomial Regression Fitting) smoothing to the search terms to capture the overall data trend, while reducing the noise introduced by weeks with zero searches. Similar approaches have been used in studies using digital data for disease modeling ^49,50^. The LOESS parameter was set at 0.045.

The autoregressive models were used as our baseline models. In the first model, ILI at time t was modeled using ILI data from time t-1. In the second model, ILI at time t was modeled using ILI data from time t-1 and a time variable. Lastly, ILI at t was modeled using ILI data from time t-1 and t-2.

## Data Availability

The Google data is publicly available.

## ACKNOWLEDGMENTS

We thank Hailun Wang and Reese Sy for their contributions to the project.

## Author Contributions

E.O.N. designed the study and analyzed the data. All authors interpreted the results. E.O.N., O.O. and M.L.M. wrote the manuscript. All authors edited the manuscript. ASAA provided data.

